# Analysis of Alzheimer’s Disease–Related Alterations in EEG Dynamics Using Integrated Instantaneous Frequency–Amplitude Microstates

**DOI:** 10.64898/2026.03.10.26347997

**Authors:** Sou Nobukawa, Takashi Ikeda, Mitsuru Kikuchi, Tetsuya Takahashi

## Abstract

Disruptions in large-scale electroencephalography dynamics are a hallmark of Alzheimer’s disease. However, conventional microstate analyses rely primarily on amplitude-based features and may overlook phase-related alterations in network organization. This study examined whether integrating instantaneous frequency and instantaneous amplitude into a unified microstate framework could better characterize AD-related EEG dynamics. Resting-state electroencephalography data were recorded from 16 patients with Alzheimer’s disease and 18 healthy controls using 16 scalp electrodes. Instantaneous frequency and instantaneous amplitude were derived via the Hilbert transformation in the theta to alpha band, ranging from 4 to 13 Hz, spatially normalized, and jointly clustered using the *k*-means algorithm with *k* equal to 4 to define the integrated frequency and amplitude microstates. Temporal properties, including dwell time, fractional occurrence, and transition probabilities, were compared between groups. The analysis identified recurrent instantaneous frequency and instantaneous amplitude microstates. Patients with Alzheimer’s disease showed a reduced occurrence of the occipital-leading state with frontal amplitude enhancement and an increased occurrence of the frontal-leading, frontal-amplified state, while transition probabilities did not differ significantly. These findings suggest that impairments related to Alzheimer’s disease are reflected in the altered prevalence of integrated phase and amplitude brain states, supporting integrated instantaneous frequency and instantaneous amplitude microstates as a complementary approach based on electroencephalography for probing neurodegenerative network dysfunction.

## I. Introduction

The emergence of diverse and sophisticated brain functions stems from the complex, coordinated operation of neural networks that span multiple cortical and subcortical regions (reviewed in [1]–[4]). Importantly, this high-dimensional neural activity is inherently dynamic, characterized by spontaneous temporal transitions during the resting state, even in the absence of external stimuli (reviewed in [5]). Characterizing these spontaneous large-scale neural dynamics is fundamental to understanding both normal cognitive processes and pathological alterations. Electroencephalography (EEG) microstate analysis represents a well-established approach for quantifying these transient brain states [6], [7] (reviewed in [8]). This method identifies quasi-stable topographical patterns that are traditionally derived from the spatial distribution of EEG oscillatory power specifically amplitude envelopes. The temporal properties and transition dynamics of these power-based microstates have proven to be valuable for investigating large-scale functional networks, providing insights into cognitive function as well as a wide range of neurological and psychiatric disorders [9]–[13].

However, capturing the full complexity of large-scale brain network dynamics necessitates consideration neural features beyond oscillatory power. Accumulating neuroscientific evidence has highlighted the critical role of phase-related characteristics in mediating interregional communication and information transfer (reviewed in [14]–[18]). Phase-based mechanisms—including phase synchronization, widely used in functional connectivity analysis [15] and cross-frequency interactions, such as phase–amplitude coupling [16]—play essential roles in coordinating neural communication across distributed brain regions. Motivated by these findings, our group previously developed a dynamic phase synchronization (DPS) metric that successfully identifies age-related changes in neural network dynamics based on temporal patterns of phase differences [19]. Subsequently, we demonstrated that the spatial organization defined by the instantaneous frequency (IF) of neural activity resembles conventional amplitude-based microstates while exhibiting distinct transition dynamics [20]. This observation led to the introduction of the IF microstate framework, a novel approach grounded in instantaneous phase characteristics. The dynamic properties of IF microstates are sensitive to the pathological alterations associated with cognitive decline in Alzheimer’s disease (AD) [21] and schizophrenia [22]; this indicates that IF microstates provide complementary information to the conventional microstate framework [23].

Impairment in the integration of neural activity is a hallmark of various pathological conditions, particularly those associated with cognitive decline (reviewed in [24]–[26]). Driven by the global increase in life expectancy, the prevalence of AD—the most common form of dementia—is projected to increase substantially, reaching approximately 1.2% by 2046 [27]. Although therapeutic strategies remain actively debated, the importance of early diagnosis and intervention to slow disease progression has been increasingly recognized [28]. Consequently, the development of reliable biomarkers for early detection has become critical. AD pathology, characterized by progressive neuronal loss and accumulation of neurofibrillary tangles and senile plaques, leads to cognitive dysfunction through the disruption of distributed neural interactions [29]– [31]. In line with this perspective, previous EEG studies have demonstrated that AD progression is reflected in alterations of frequency-band-specific global functional connectivity [32], [33]. Moreover, numerous studies investigating whole-brain network dynamics—including dynamic functional connectivity analyses using sliding time windows [34]–[36] and power topography based on microstate analyses [37]–[39]—have consistently reported altered network dynamics in AD. This is further supported by our IF microstate analysis [21].

Despite growing evidence that both phase- and amplitude-related dynamics contribute to large-scale brain network functions [40], the precise interaction between these two aspects within transient brain states—particularly in the context of neurodegenerative disorders—remains unclear. In our preliminary study, we introduced a unified microstate framework that integrates the spatial distributions of IF and instantaneous amplitude (IA)—termed IF–IA microstates—and applied it to healthy aging populations [40]. While this initial investigation demonstrated the feasibility of such integration, the relationship between the phase-leading or -lagging patterns captured by IF microstates and concurrent amplitude dynamics has not yet been examined under pathological conditions. To address this gap, we hypothesized that jointly integrating the spatial distributions of IF and IA within the IF–IA microstate framework would enable a more comprehensive characterization of spatiotemporal neural dynamics associated with cognitive impairment in AD. Accordingly, we defined IF–IA microstates using resting-state EEG data recorded from patients with AD and healthy controls (HC). By analyzing the dynamic properties of these IF–IA microstates, we aimed to identify AD-specific neural signatures and evaluate the potential of the IF–IA microstate framework as a complementary EEG-based biomarker of neurodegenerative network dysfunction.

## II. Materials and methods

### A. Participants

In this study, we analyzed EEG data acquired from two distinct cohorts: patients diagnosed with AD and older HC adults. The AD group consisted of 16 individuals who met the National Institute of Neurological and Communicative Disorders and Stroke-Alzheimer’s Disease and Related Disorders Association criteria and were in the pre-dementia phase according to the Diagnostic and Statistical Manual of Mental Disorders (fourth edition) criteria. The HC group comprised sex- and age-matched older participants who were non-smokers and were not taking any medications at the time of the experiment. Exclusion criteria for the HC group included any history of major medical or neurological conditions— such as epilepsy or severe head trauma—and a history of substance (alcohol or drug) dependence. Care was taken to ensure that no medications affecting the central nervous system were administered to patients with AD during the study period. All participants with AD group underwent comprehensive cognitive and functional assessments using the Functional Assessment Staging Test (FAST) and the Mini-Mental State Examination (MMSE) [41]. The severity distribution within the AD cohort included 3 participants with mild AD (FAST 3), 7 with moderate AD (FAST 4), and 6 with severe dementia (FAST 5), with MMSE scores ranging from 10 to 26 (mean = 15.56). Detailed demographic and clinical characteristics of all participants are summarized in Table I. All participants provided written informed consent prior to the inclusion in the study. The experimental protocol was approved by the Ethics Committee of Kanazawa University and conducted in strict accordance with the Declaration of Helsinki.

**TABLE I.** Demographic and clinical characteristics of healthy controls (hcs) and patients with alzheimer’s disease (ad). data are presented as mean (standard deviation, range) or frequency.

|  | HC participants | AD participants | *p* values |
| --- | --- | --- | --- |
| Male/female | 7/11 | 5/11 | 0.72 |
| Age (years) | 59.3 (5.3, 55–66) | 57.5 (4.7, 43–64) | 0.31 |
| MMSE score | NA | 15.5 (4.7, 10–26) | NA |

### B. EEG Recording and Preprocessing

Resting-state EEG signals were recorded with eyes closed for a duration of 10 to 15 minutes. To minimize external interference, recordings were conducted in a dedicated, electrically shielded, and soundproof room with controlled lighting. A 16-channel EEG system (EEG-4518, Nihon Kohden, Tokyo, Japan) was used. Sixteen scalp electrodes were positioned according to the international 10-20 system at the following locations: Fp1, Fp2, F3, F4, C3, C4, P3, P4, O1, O2, F7, F8, Fz, Pz, T5, and T6. All signals were referenced to a binaural connection. The technical specifications for recording were as follows: Sampling Frequency, 200 Hz; Online Bandpass Filter, 1.5–60 Hz (time constant, 0.3 s). The electrode–skin impedance was strictly maintained below 5kΩ at all recording sites. Eye movements, monitored via bipolar electrooculography, were continuously traced. For the subsequent analysis, a continuous, artifact-free epoch of 60 s (12, 000 data points) was carefully selected and extracted from the full recording period for each participant to ensure data quality and comparability across subjects.

### C. Estimation of IF-IA Microstates

The characterization of large-scale neural states in this study relied on the integration of the IF and IA dynamics derived from EEG signals. The overall procedure is visually summarized in (Fig. 1). Initially, the multichannel EEG recordings underwent bandpass filtering to isolate the 4–13 Hz thetaalpha band, which is known to be the dominant oscillatory regime during the eyes-closed resting state. Subsequently, a Hilbert transform was applied to the filtered signals to obtain an analytical signal. From this, the wrapped instantaneous phase *θ*(*t*) constrained to the range (*−π ≤ θ ≤ π*) and the IA time series were extracted. To mitigate potential inaccuracies in the IF estimation caused by phase noise, specifically phase slips [42], a median filter was applied to both the derived IF and IA time series. This filtering approach follows established practices used in our previous study on DPS analysis [19].

**Fig. 1.**
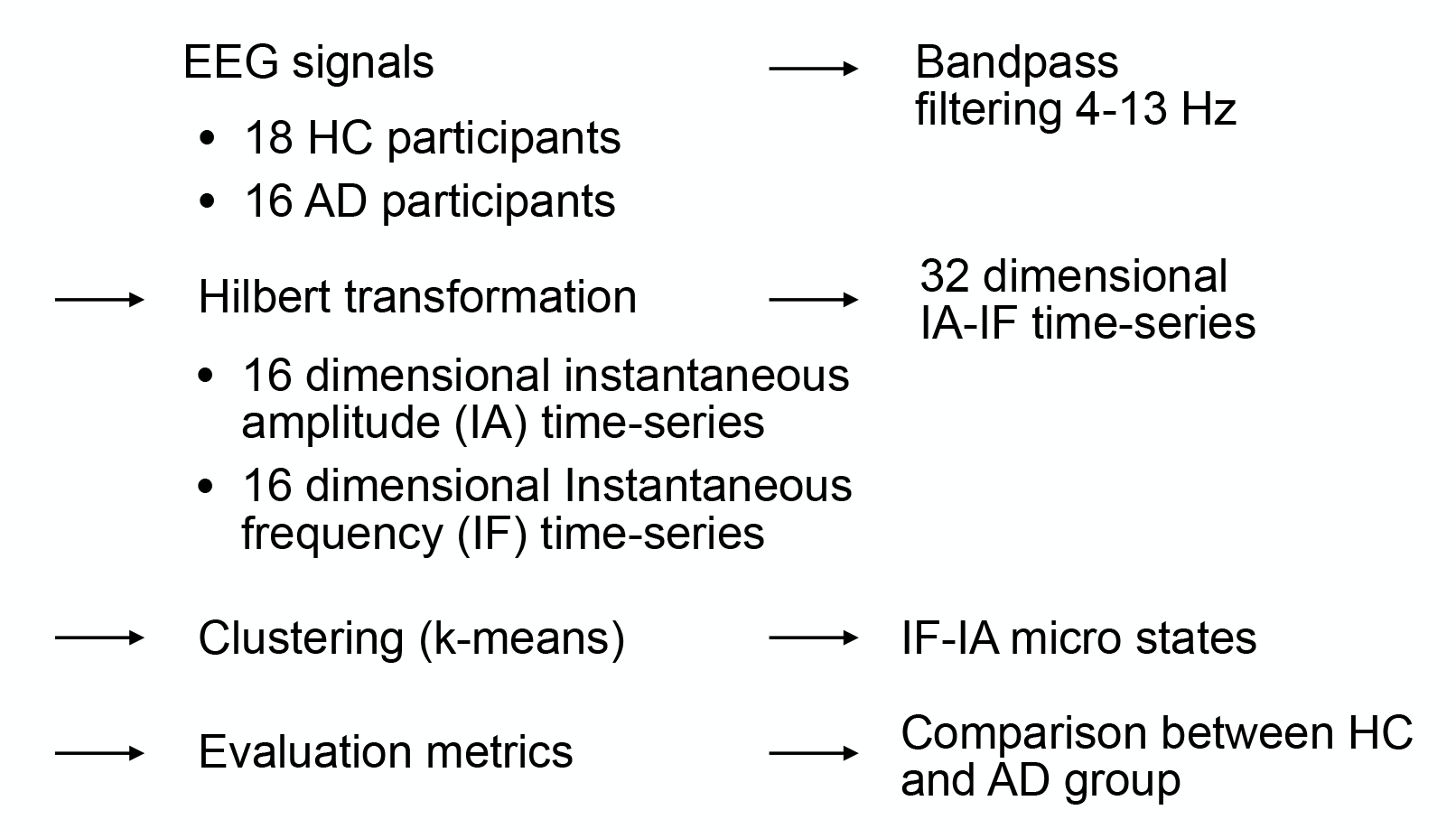
Overview of the estimation methodology for IF-IA microstates in Alzheimer’s disease (AD) and healthy control (HC) groups. This process involves the derivation of instantaneous frequency (IF) and instantaneous amplitude (IA) time series from the multichannel electroencephalography (EEG), followed by spatial normalization and subsequent clustering to identify recurring global brain states.

#### 1) Spatial Deviation and Normalization

To define the spatial brain state at any moment, the spatial deviations of the IF and IA were computed for each electrode *i*. This process involves centering the activity by subtracting the average across all electrodes at time *t*. The instantaneous frequency deviation *dIF*_*i*_(*t*) is calculated as follows:

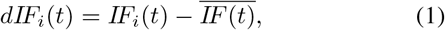

where, 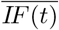 represents the mean IF across all the electrodes at time *t*. Similarly, the instantaneous amplitude deviation *dIA*_*i*_(*t*) is computed by centering the IA signal

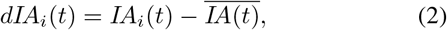

where, 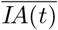 denotes the mean IA across all the electrodes at time *t*. Both *dIF*_*i*_(*t*) and *dIA*_*i*_(*t*) time series were subsequently *z*-scored at each time point to ensure that the analysis focused on relative spatial patterns across the scalp rather than absolute values.

#### 2) Microstate Classification

Each time point *t* was represented as a 32-dimensional feature vector (16 electrodes *×* two features: *dIF*_*i*_ and *dIA*_*i*_). To identify recurring quasistable spatial patterns, the *k*-means clustering algorithm was applied to the pooled data from both patients with AD and HC participants. The number of clusters is set to *k* = 4, which is consistent with our previous study [40]. The resulting four cluster maps defined the IF–IA microstates, representing rapid reconfigurations of large-scale neural activity. Thus, the temporal sequence of cluster labels describes the moment-to-moment evolution of brain states.

### D. Evaluation Indexes

#### 1) Temporal and Dynamic Metrics

To quantify the temporal characteristics and dynamic flexibility of the identified IF-IA microstates, we employed several standard evaluation metrics. For each distinct microstate *i*, we calculated the temporal indices as follows: The dwell time (*D*_*i*_) is defined as the mean duration of the microstate’s continuous and uninterrupted appearances. The calculation is given by

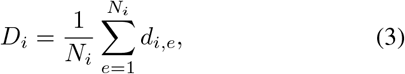

where *N*_*i*_ denotes the total count of microstate *i*’s occurrences, and *d*_*i,e*_ represents the temporal length of the *e*-th episode. The occurrence ratio (*f*_*i*_) is a metric that quantifies the proportion of the total analyzed time (*T*) in which microstate *i* is active. The formulation is:

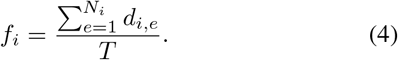

We characterized the dynamic switching behavior between the identified microstates by calculating the state transition probabilities *P* (*i → j*) for all the possible ordered pairs (*i → j*). The probability of transitioning from state *i* to state *j* is determined by the ratio of observed transitions to the total number of exits from state *i*:

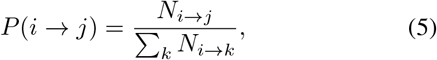

where *N*_*i→j*_ is the measured frequency of the transitions observed from state *i* directly to state *j*.

### E. Statistical Analysis

We conducted statistical comparisons between the AD and HC groups to determine significant differences in the dynamic properties of the IF-IA microstates. To compare microstate emergence metrics, specifically dwell time and occurrence ratio, between the HC and AD cohorts, all data underwent an initial log-transformation to effectively reduce skewness before hypothesis testing. The distribution of the transformed data was subsequently assessed for normality using the D’Agostino–Pearson test. Group differences were evaluated using a two-sample *t*-test when data from both groups satisfied the normality criteria; otherwise, the nonparametric Mann– Whitney *U* test was employed. To control for the family wise error rate stemming from multiple comparisons across the four identified microstates, the results were subjected to a False Discovery Rate (FDR) correction, with statistical significance established at a corrected threshold of *q <* 0.05.

For the transition probabilities, statistical assessments were performed for every possible transition, totaling *k*^2^ = 16 pairs between four microstates. The selection of the appropriate test (*t*-test or Mann–Whitney *U* test) followed the same normality assessment procedure as that employed for the temporal metrics. Consistent with other analyses, FDR correction was applied globally across all 16 calculated transition probabilities to ensure the robustness of our statistical inferences.

## III. Results

First, we evaluated the spatial patterns of IF and IA corresponding to the four identified IF–IA microstates. Figure 2 presents the group-averaged *dIF*_*i*_ and *dIA*_*i*_ distributions in AD and HC participants. State #2 exhibited a clear occipital-leading pattern in IF, accompanied by increased IA over the frontal regions. State #4 exhibited frontal-leading and frontal-amplified patterns. By contrast, State #1 presented a relatively ambiguous leading pattern with only left-lateralized amplitude modulation. State #3 shows a relatively ambiguous leading pattern, characterized by a right-lateralized occipital amplification pattern.

**Fig. 2.**
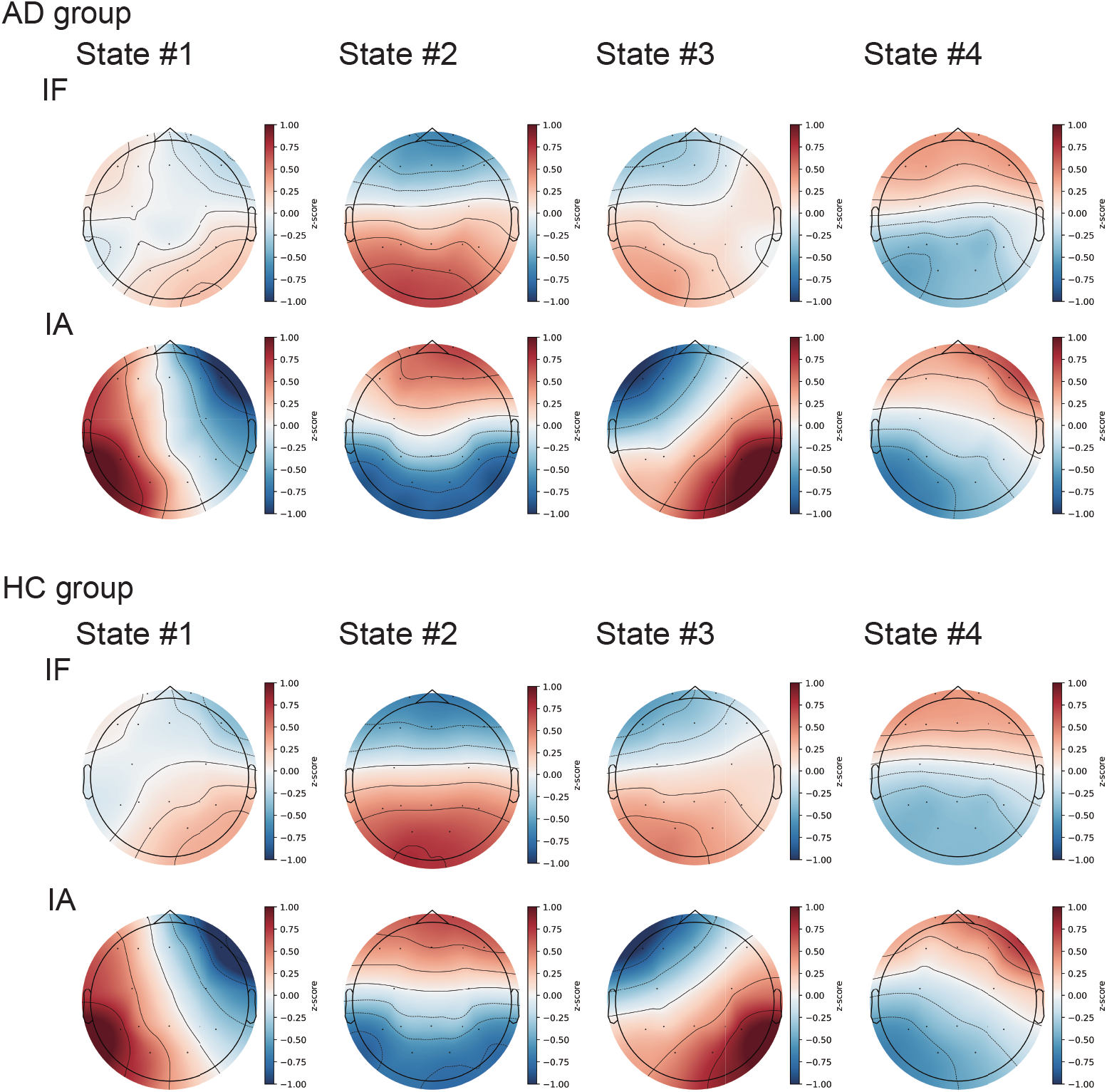
Group-averaged spatial distributions of instantaneous frequency (*dIF*) and instantaneous amplitude (*dIA*) for the four IF–IA microstates in the AD and HC groups. State #2 demonstrates an occipital-leading IF pattern with frontal amplitude enhancement, whereas State #4 is characterized by frontal leading and frontal-dominant amplitude enhancement. States #1 and #3 exhibit relatively ambiguous leading patterns, accompanied by left- and right-lateralized occipital amplitude enhancement, respectively.

To evaluate the dynamic properties of the IF-IA microstates, the dwell time *D*_*i*_ and occurrence ratio *f*_*i*_ were compared between the AD and HC groups, as shown in Fig. 3 (A). The occurrence ratio *f*_*i*_ for states #2 and #4 showed a significant reduction and increase, respectively, compared to the HC group (*q <* 0.05). Moreover, to evaluate the transition characteristics among IF-IA microstates, the transition matrix for the AD and HC groups and signed significance maps (sign *×−* log_10_ *p*) illustrating the statistical differences between the groups are shown in Fig. 3 (B). No statistically significant differences were detected in transition probabilities between the AD and HC groups.

**Fig. 3.**
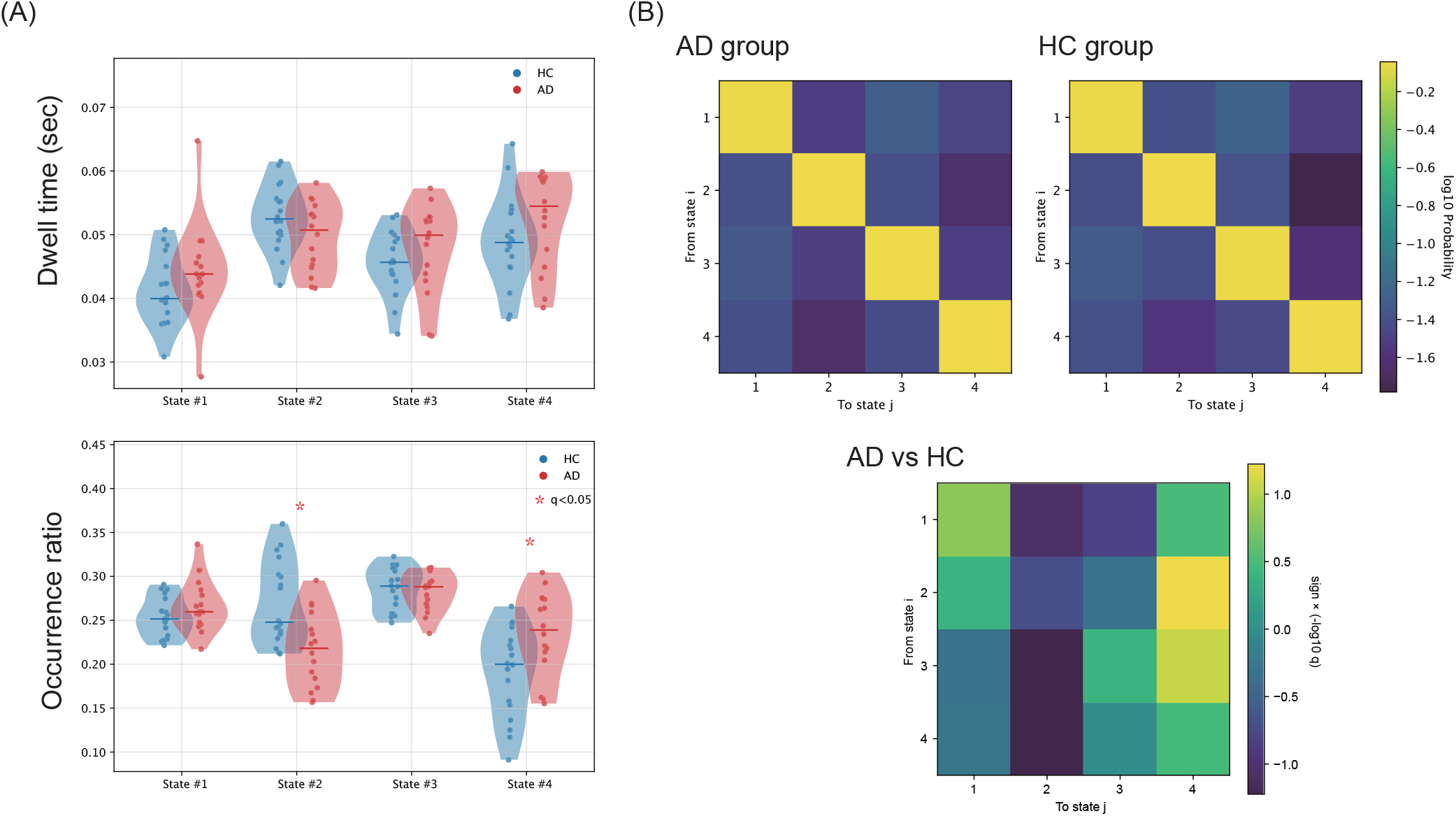
Dynamic properties of IF-IA microstates in AD and HC groups. (A) Dwell time *D*_*i*_ (upper panel) and occurrence ratio *f*_*i*_ (lower panel) for AD and HC groups. Individual participants are represented by dots (red for AD, blue for HC); horizontal lines indicate group means, and shaded violin plots visualize the distribution shape. The occurrence ratio *f*_*i*_ for State #2 and State #4 showed a significant reduction and increase, respectively, when compared to the HC group (*q <* 0.05). (B) Mean transition probabilities among IF-IA microstates. The transition matrices for the AD group (upper left) and the HC group (upper right). The lower panel displays Signed Significance Maps (sign *× −* log_10_ *p*), illustrating the statistical differences between the groups. Positive (negative) values indicate higher (lower) transition probabilities in the AD group compared to the HC group. Note that no statistically significant differences were detected in transition probabilities between the AD and HC groups.

## IV. Discussion

This study investigated whether integrating IF and IA into a unified microstate framework could more comprehensively characterize the large-scale EEG dynamics associated with AD. To this end, IF–IA microstates were defined by jointly clustering the spatial deviations of IF and IA derived from resting-state EEG in patients with AD and healthy controls. The dynamic properties of the resulting microstates were systematically evaluated using standard microstate metrics, including dwell time, occurrence ratio, and state transition probabilities. The analysis identified four recurrent IF–IA microstates, among which patients with AD exhibited a significantly reduced occurrence of an occipital-leading state with frontal amplitude enhancement and an increased occurrence of a frontal-leading, frontal-amplified state, whereas no significant group differences were observed in transition probabilities. These findings indicate that AD-related alterations are primarily reflected in the relative prevalence of specific integrated phase–amplitude brain states, rather than in the overall switching structure, highlighting the utility of IF–IA microstates in capturing disease-related impairments in large-scale neural dynamics.

Based on these results, the mechanisms underlying the reduced occurrence of the occipital-leading state with frontal amplitude enhancement and the increased occurrence of the frontal-leading and frontal-amplified states in AD (see Fig. 3 (A)) should be considered. In our previous study, in which AD EEG signals were analyzed using IF microstates without incorporating IA information [21], we observed a reduced appearance of occipital-leading IF microstates. The pathological progression of AD impairs the posterior cingulate gyrus, which plays a critical role as a hub for large-scale brain networks involved in information integration and transmission (reviewed in [43]–[45]). Assuming that phase-leading states reflect neural transmission processes [46], the dysfunction of this hub region, which is associated with cognitive decline in AD [47]–[49] may contribute to difficulties in generating occipital-leading states. Because of this reduced posterior-driven transmission, frontal-leading states may appear more frequently, reflecting a shift in the dominant patterns of large-scale neural dynamics. Notably, the reduction in occipital-leading states observed in this study was specific to states accompanied by frontal amplitude enhancement. This finding suggests that large-scale network integration and transmission mediated by posterior hub regions, such as the posterior cingulate gyrus, may have been originally implemented in an occipital-leading state with concomitant frontal amplification. Accordingly, the pathological disruption of the posterior hub function in AD may render this integrated state difficult to sustain, resulting in the selective reduction of occipital-leading states under frontal-amplified conditions.

Several limitations of this study warrant consideration. First, the number of IF–IA microstates was predetermined at *k* = 4 when the clustering procedure was applied to the instantaneous spatial patterns of IF and IA. While this selection was informed by conventional criteria, identifying the optimal cluster number to comprehensively capture the diversity of IF–IA microstates remains an open challenge that warrants further systematic investigation. Second, prior research has established that alterations in large-scale network dynamics are not exclusive to AD but are also manifest across a broad spectrum of psychiatric conditions [50], [51]. Accordingly, distinct disorder-specific signatures may reside within IF–IA microstate dynamics—a possibility that was not explored in the current study. Future investigations encompassing a more diverse clinical spectrum and rigorous analytical validation are essential to further elucidate these phenomena.

## V. Conclusions

In conclusion, this study establishes that IF–IA microstates, which synthesize instantaneous frequency and amplitude information, provide a robust framework for characterizing alterations in large-scale EEG dynamics associated with AD. The observed shifts in the relative prevalence of specific IF– IA microstates suggest that AD-related network impairment is manifested as a disruption of integrated phase–amplitude brain states, rather than a deviation in global state-switching patterns. Specifically, the selective reduction of occipital-leading states under frontal-amplified conditions likely underscores the vulnerability of posterior hub–mediated integration processes in AD. These findings indicate that the IF–IA microstate framework offers a sophisticated complementary perspective to conventional microstate analyses and holds significant potential for the development of reliable EEG-based biomarkers for neurodegenerative disorders.

## Data Availability

The datasets generated for this study will not be made publicly available, because informed patient consent will not include a declaration regarding the public availability of the clinical data.

## Acknowledgment

The authors declare that this study was conducted in the absence of any commercial or financial relationships that could be construed as conflicts of interest. The experimental protocol of this study was approved by the Ethics Committee of Kanazawa University and conducted in accordance with the Declaration of Helsinki. All participants provided written informed consent to participate in the study. Portions of the text, including language refinement and editing, were generated using ChatGPT-5, an artificial intelligence language model developed by OpenAI.

